# Prevalence and associated factors of antenatal depression in rural Bangladesh

**DOI:** 10.1101/2024.05.30.24308225

**Authors:** Rifa Tamanna Mumu, Dipak Kumar Mitra

**Affiliations:** Department of Public Health, School of Health and Life Science, North South University, Bashundhara, Dhaka, Bangladesh

**Keywords:** Antenatal Depression, Mental Health in Pregnancy, Rural Bangladesh, Factors influencing maternal mental health, Prenatal depression, Prevalence, Associated Factors, Risk Factors, Depression

## Abstract

**Background:** According to the World Health Organization (WHO), approximately 322 million individuals globally were grappling with depressive disorders in 2015. During pregnancy, the risk of experiencing depression is elevated due to certain hormonal changes. Despite the potentially severe consequences of antenatal depression for both the mother and newborn, there have been limited studies conducted on this issue in Bangladesh.

**Objective:** To find out the prevalence and associated factors of antenatal depression in a rural sub-district in Bangladesh.

**Method:** A cross-sectional study was performed in Lohagara, a rural subdistrict in Narail, situated in the southern part of Bangladesh between January 08 and 14, 2024. 350 subjects were recruited for the study, who were pregnant at various trimesters and attended antenatal check-ups in a government health complex and a private hospital in Lohagara. The Bengali-translated version of the Edinburgh Postnatal Depression Scale (EPDS) and a structured questionnaire were used for data collection. Data were analyzed in STATA version 14.

**Result:** The point prevalence of antenatal depression is 39% (38.86%, in 95% CI: 33.9% to 44%). Gestational week (AOR: 0.4, 95% CI: 0.2, 0.8), unintended pregnancy (AOR: 1.7, 95% CI: 1, 3), intimate partner violence (AOR: 3.3, 95% CI: 1.1, 9.7), a history of previous diseases (AOR: 2.4, 95% CI: 1.1, 5.2), and the history of having polygamous husbands (AOR: 13.6, 95% CI: 1.1, 164) are found to be significantly associated with the development of depression in the prenatal period.

**Conclusion:** In rural Narail, high rates of antenatal depression underscore the importance of increased awareness among healthcare professionals and families. Strategic involvement of stakeholders and policymakers is essential to address issues like intimate partner violence and polygamy. Moreover, there’s a critical need for extra care and counseling for pregnant women with a history of health problems or facing unexpected pregnancies.

## Introduction

Globally, depression stands out as a prevalent mental health disorder, marked by symptoms such as a low mood, changes in appetite and sleep patterns, loss of interest in activities, significant weight fluctuations, feelings of hopelessness, concentration difficulties, low self-esteem, and frequent thoughts of mortality. It holds a prominent position among the top five causes of the global disease burden (1). Projections suggest that by 2030, depressive disorders will likely become one of the three leading contributors to the overall global burden of disease (2).

In 2015, the World Health Organization (WHO) approximated that there were 322 million individuals globally experiencing depressive disorders and 27% of them were from the Southeast Asian region (1). The risk of mental disorders, especially depression is more in females than in males (3). In pregnancy, the risk of depression is higher than general female population due to hormonal changes (4). The global occurrence of antenatal depression ranges from 15% to 65% (5). In high-income countries, the prevalence ranges from 5% to 30% (6, 7, 8), while in low-income countries, it is 15.6% to 31.1% (9, 10, 11).

The occurrence of depression during pregnancy in Bangladesh varies between 18% and 33% (12), which is not too small. A recent study, carried out in a rural sub-district in Matlab, involving pregnant women at their 34-35 weeks of pregnancy showed a 33% prevalence and the associated factors included an unsupportive husband or mother-in-law, domestic violence, and mental pressure for male gender preference of the family (13). A recent cross-sectional study, conducted in rural Sylhet highlighted male gender preference of husband, low family support, and sexual violence as the main associated factors for developing antenatal depression (12).

The consequences of depression in pregnancy can be devastating and it may affect both mother and child. A pregnant woman experiencing depression may release a hormone called cortisol, which can have detrimental effects on fetal growth and brain development (14). Women with antenatal depression have a higher risk of developing Hyperemesis gravidarum, which increases the probability of miscarriage, low birth weight, and preterm birth (15). Not only that, the risk of substance abuse, preeclampsia, hemorrhage, edema, postpartum depression, and severe headaches is higher in a woman with antenatal depression (16, 17). The neonatal outcomes of antenatal depression are reported as low birth weight (LBW), low mean APGAR scores at 1 and 5 minutes following birth, and premature mortality (18, 19).

Antenatal depression poses a significant challenge for expectant mothers, particularly in Bangladesh. Research on this topic is predominantly limited to rural areas, with scant urban-based studies available. Moreover, there is a notable dearth of information regarding the prevalence of depression during different trimesters of pregnancy in Bangladesh. This gap in research underscores the need for a more comprehensive understanding of antenatal depression across diverse geographic and demographic settings in the country. The absence of research on antenatal depression in Narail, a southern district in Bangladesh, underscores the significance of this study. By shedding light on the current status of antenatal depression at the sub-district level in Narail, this research aims to provide a crucial snapshot of the situation. The findings from this study can play a pivotal role in identifying necessary interventions, informing policy-making decisions, and structuring health education programs. The ultimate goal is to enhance awareness and establish effective measures for controlling antenatal depression in the future.

## Method

### Study Design and Setting

A cross-sectional study was performed between January 08 and 14, 2024 in Upazila Health Complex, Lohagara, a government hospital, and Khan General Hospital, Lahuria, a private hospital in Lohagara, a rural sub-district in Narail, situated in the southern part of Bangladesh.

### Study Participants

The target population was pregnant mothers of any trimester in the Lohagara sub-district and the sample population was pregnant mothers of any trimesters attending the ANC Corner of Upazila Health Complex, Lohagara, Narail, and Khan General Hospital, Lahuria for antenatal checkups.

### Sample Size and Sampling Technique

Considering the prevalence of antenatal depression in Bangladesh is 33% according to a study conducted in a rural sub-district in Matlab (13), 95% confidence interval, and with 5% margin of error, calculated sample size, 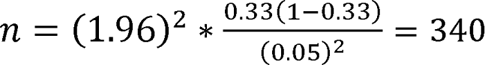

The sampling technique was systematic sampling. Every third patient attending ANC Corner for antenatal checkups in both government and private hospitals was selected as a participant in the interview for data collection.

### Data Collection Tools

The presence of depression was assessed by the Bengali-translated version of the Edinburgh Postnatal Depression Scale (EPDS). This questionnaire consists of 10 questions scoring from 0 to 30. A score of 10 or higher on the assessment indicates probable antenatal depression.

Another structured questionnaire, which was also translated into Bengali, was used to collect data on the sociodemographic, obstetric, psychosocial, psychological, and disease and treatment-related factors of patients. The questionnaire was tested on some target population rather than the study participants and the necessary changes were made before the data collection.

### Data Management & Analysis Plan

The data for the study was analyzed by STATA version 14. Pearson’s chi-square test was performed to find out the possible association of sociodemographic, obstetric, psychosocial, disease, and treatment-related factors with prenatal depression. A binary logistic regression was also performed to find out the crude odds ratio of variables. To adjust the confounding factors, a multivariate analysis using multivariate logistic regression was performed. Adjusted and unadjusted odds ratio and their 95% CI were used as indicators strength of the association.

### Ethical Considerations

Ethical permission was taken from the Institutional Ethics Committee of North South University before data collection (Approval Number: 2023/OR-NSU/IRB/1224). Permission letters from Upazila Health Complex, Lohagara, and Khan General Hospital, Lahuria were also obtained. Informed written consent was obtained from pregnant mothers of 18 years or more and guardians of pregnant women aged below 18 years before data collection. The respondents were assured of the confidentiality of information and also informed about the purpose, advantages, and potential risks of the study.

## Result

### Socio-demographic Characteristics of Pregnant Mothers

In this research involving 350 women in any trimester of gestation, with a high response rate of 98.5%, several demographic characteristics were examined. The median Edinburgh Postnatal Depression Scale (EPDS) score was 8, with an interquartile range of 4 to 12. The respondents had a median age of 23 years, falling within an interquartile range of 20 to 27 years. Among the respondents, 5.7% (20) were below 18 years old, while a majority of 83.7% were in the age bracket of 18 to 30 years. The rest 10.6% were over their thirties.

Regarding educational background, 26 participants (7.4%) did not have a minimum primary level education. All participants were married, with the majority being housewives. However, a small percentage (4%) were employed. In terms of monthly family income, 26% of women (91) had an income below 10,000 tk, 58% (203) had an income between 10,000 to 20,000 tk, and the remaining 16% (56) had an earning per month exceeding 20,000 tk.

Religiously, the participants belonged to two communities, with 96.3% (337) identifying as Muslim and 3.71% (13) as Hindu. These demographic details provide a comprehensive snapshot of the participants in the study, offering insights into their socioeconomic and cultural backgrounds.

### Obstetric History of Pregnant Mothers

A large percentage (53.4%) of respondents were in their second trimesters. 107 women (30.6%) were in the third trimester, and the rest 16% (56) were passing the first trimester. In this study, the median age of marriage was found to be 18 years, with an interquartile range spanning from 16 to 19 years. Significantly, almost half of the participants (48.6%) reported a history of getting married before the age of 18. Regarding pregnancy status, 38.6% (135) of the total expectant mothers were experiencing pregnancy for the first time.

Among the multiparous women, 51% (105) had undergone at least one cesarean section (46.6% only caesarian section, 4.4 % had both vaginal delivery and caesarian section), and 30.6% (63) faced complications during their previous deliveries. Additionally, 21.5% (75) of the women reported a history of abortion, stillbirth, or intrauterine fetal death. These findings provide important contextual information about the participants’ marital and reproductive histories, contributing to a more comprehensive understanding of the factors influencing antenatal depression in this population.

### Psychosocial Criteria of Pregnant Mothers

In the study, 74% of the women (259) reported that their current pregnancies were planned, while 26% (91) indicated the opposite. Regarding satisfaction with their husbands’ behavior, the majority of women (50.3%) expressed moderate satisfaction, while a small proportion (4.8%) reported poor relationship status.

Concerning relationships with in-laws, the majority (77.8%) reported good relationships, and 15.7% (55) described their relationships as moderate. However, 6% (21) of women had a history of poor relationship status, with 1.43% (5) being separated from or having deceased parents-in-law.

The study also highlighted instances of violence. Approximately 5.14% of women (19) reported being victims of domestic violence, and 5.43% (18) reported experiencing sexual violence. Notably, 10% of women (35) had faced sexual violations during the current pregnancy.

Additionally, family dynamics played a role, with 18.6% of women (65) reporting that their husbands preferred male children, and 18% (63) mentioned specific demands from their in-laws for a male child. These findings provide insights into the social and familial factors that may contribute to antenatal depression in the study population.

### Disease and Treatment-related History of Pregnant Mothers

Among the 350 women, 37 (10.57%) were suffering from diseases like Diabetes Mellitus, Hypertension, Bronchial Asthma, and Thyroid disorders, while 49 (14%) had a history of at least one previous surgery other than caesarian section.

### Psychological Factors of Pregnant Mothers

In the study, a small percentage of women reported certain marital challenges. Specifically, 2.57% (9) of the women mentioned that their husbands had other wives, and a couple of women (0.6%) indicated a history of their husbands having extra-marital relationships with other women. However, none of the participants reported having such relationships themselves. These findings offer insights into the complex dynamics within marital relationships and potential factors that may contribute to antenatal depression in the studied population.

### Prevalence of Depression among Pregnant Mothers

In the study, the scores from the 30 questions of the Edinburgh Postnatal Depression Scale (EPDS) were summed up, creating a new variable ranging from 0 to 30. The findings revealed that 150 (42.9%) women had none or minimal depression, and 141 women (40.29%) had mild depression, scoring between 7 to 13 on the EPDS. Another 43 women (12.29%) scored between 14 to 19, indicating moderate depression. Additionally, 16 women (4.57%) had severe depression, with scores exceeding 19. Table 1

**Table 1.**
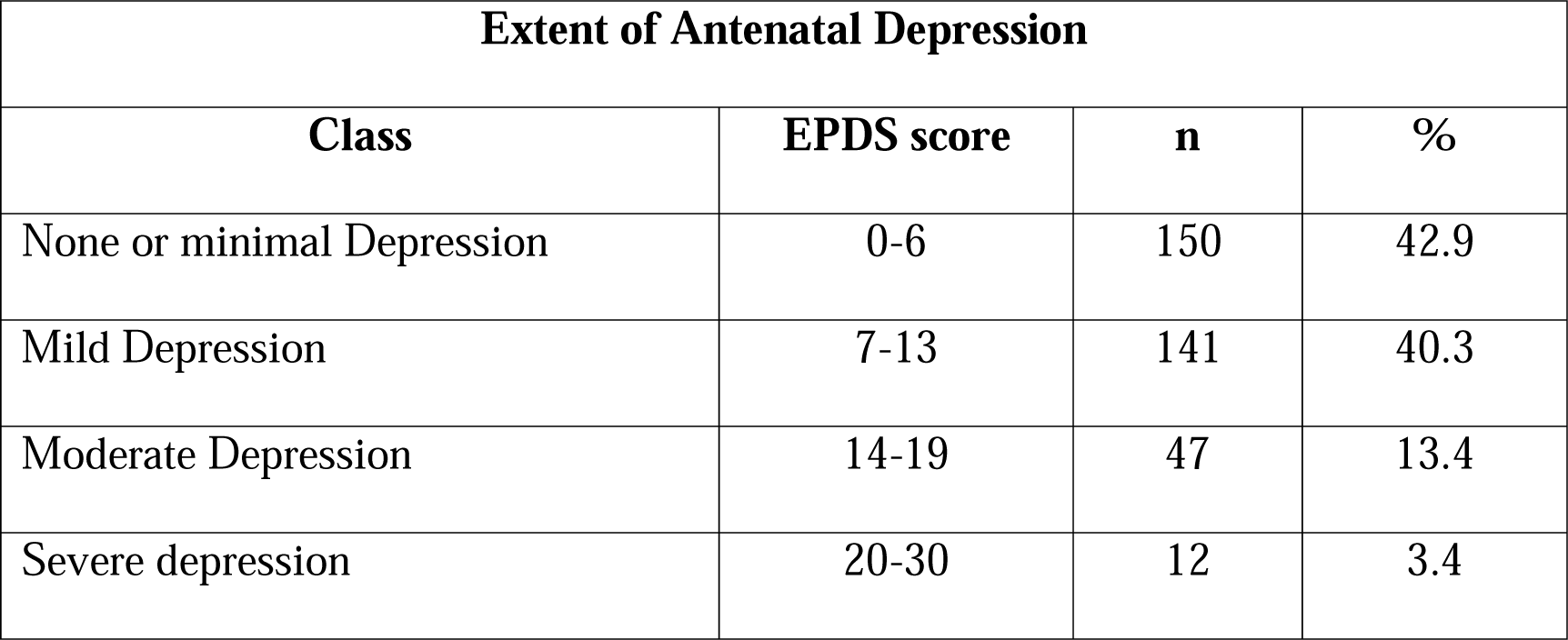
Extent of Depression among pregnant women in Lohagara.

**Table 2.**
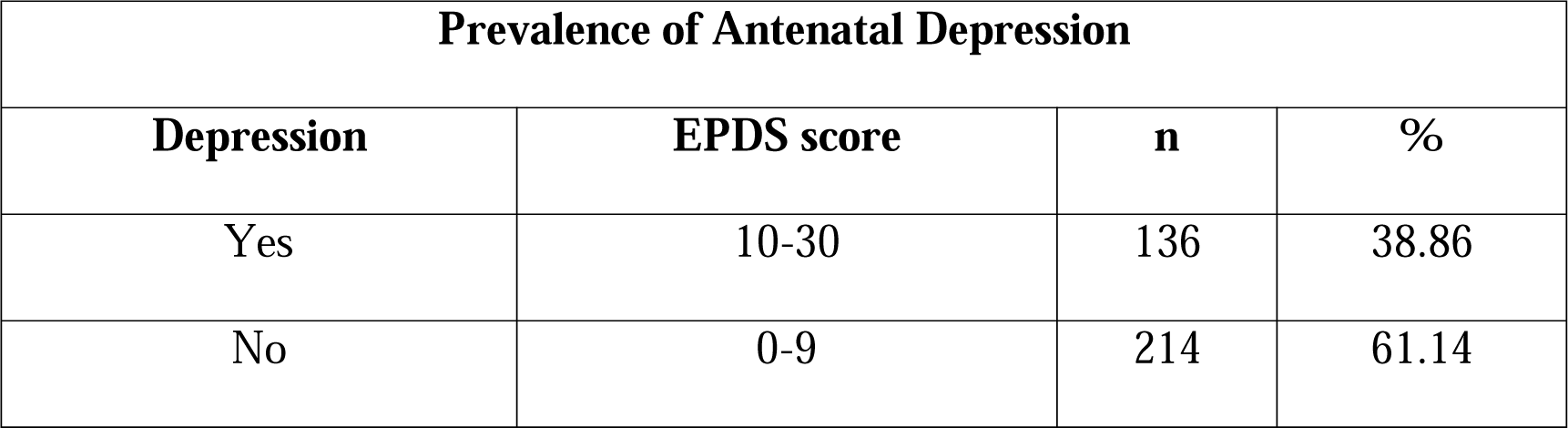
Prevalence of Antenatal Depression in Lohagara.

| Prevalence of Antenatal Depression |  |  |  |
| --- | --- | --- | --- |
| Depression | EPDS score | n | % |
| Yes | 10-30 | 136 | 38.86 |
| No | 0-9 | 214 | 61.14 |

To simplify the interpretation, the scores were further categorized into two classes: “Having Depression” and “No Depression.” A cut-off score of 10 or more was used to indicate the presence of depression. The results showed that nearly one in three women (38.86%, with a 95% confidence interval ranging from 33.9% to 44%) exhibited symptoms of depression during pregnancy. These figures highlight the prevalence and varying degrees of depression among the study participants **Error! Reference source not found.**.

### Factors associated with Antenatal Depression

Sociodemographic, Obstetric, Psychosocial, Psychological, and Disease and treatment-related factors were used to identify the factors that were significantly associated with the development of depression in pregnancy. Among the variables, gestational week, number of pregnancies, type of pregnancy, relationship with husband as well as in-laws, history of domestic and sexual violence, having a husband with multiple marriages, and history of previous disease were found statistically significant with a p-value <0.05 in the chi-square test Table 3.

**Table 3.** Factors associated with antenatal depression in women visiting for antenatal check-ups in UHC, Lohagara, and Khan General Hospital, Lahuria from July to August 2023 (bivariate analysis by chi-squared test).

| Variables | Category | No Depression | Mild Depression | Moderate depression | Severe Depression | Chi2 p-value |
| --- | --- | --- | --- | --- | --- | --- |
| Gestational week | <12 weeks | 14<br>9.3% | 30<br>21.3% | 10<br>21.3% | 2<br>16.7% | 0.026 |
|  | 12-28 weeks | 92<br>61.3% | 74<br>52.5% | 15<br>31.9% | 6<br>50% |  |
|  | 29-40 weeks | 44<br>29.3% | 37<br>26.2% | 22<br>46.8% | 4<br>33.3% |  |
| Para | Nulliparous | 68<br>45.3% | 48<br>34.4% | 16<br>34.4% | 3<br>25% | 0.005 |
|  | Multiparous | 82<br>54.7% | 93<br>66% | 31<br>66% | 9<br>75% |  |
| Type of pregnancy | Planned | 123<br>82% | 97<br>68.8% | 35<br>72.3% | 4<br>33.3% | 0.002 |
|  | Unplanned | 27<br>18% | 44<br>21.2% | 12<br>27.7% | 8<br>66.7% |  |
| Relationship with husband | Good | 72<br>48% | 60<br>42.6% | 22<br>46.8% | 3<br>25% | 0.017 |
|  | Moderate | 73<br>48.7% | 78<br>55.3% | 20<br>42.6% | 5<br>41.7% |  |
|  | Poor | 5<br>3.3% | 3<br>2.1% | 5<br>10.6% | 4<br>33.3% |  |
| Relationship with in-laws | Good | 128<br>85.3% | 110<br>78% | 28<br>79.6% | 3<br>25% | 0.001 |
|  | Moderate | 16<br>10.7% | 24<br>17% | 12<br>25.5% | 3<br>25% |  |
|  | Poor | 5<br>3.3% | 4<br>2.8% | 7<br>14.9% | 5<br>41.7% |  |
|  | Dead/Separated | 1<br>0.7% | 3<br>2% | 0 | 1<br>8.3% |  |
| Domestic violence | Yes | 5<br>3.3% | 7<br>5% | 4<br>8.5% | 2<br>16.7% | 0.047 |
|  | No | 145<br>96.7% | 134<br>95% | 43<br>91.5% | 10<br>83.3% |  |
| Sexual violence | Yes | 4<br>2.7% | 9<br>6.4% | 3<br>6.4% | 3<br>25% | 0.007 |
|  | No | 146<br>97.3% | 132<br>93.6% | 44<br>93.6% | 9<br>75% |  |
| Polygamous husband | Yes | 1<br>0.7% | 4<br>2.8% | 1<br>2% | 3<br>25% | 0.002 |
|  | No | 149<br>99.3% | 137<br>97.2% | 46<br>97.9% | 9<br>75% |  |
| Previous disease | Yes | 9<br>6% | 18<br>12.8% | 7<br>14.9% | 3<br>25% | 0.007 |
|  | No | 141<br>94% | 123<br>87.2% | 40<br>85% | 9<br>75% |  |

A binary logistic regression was also performed to find out the odds ratio of variables Table 4. These covariates were then considered for the multiple logistic regression analysis.

**Table 4.** Factors associated with prenatal depression among women attending antenatal care in UHC, Lohagara and Khan General Hospital, Lahuria from June to August 2023 (after bivariate and multivariate regression analysis).

**(after bivariate and multivariate regression analysis).**
| Variables | Category | Depression | No | COR | AOR |
| --- | --- | --- | --- | --- | --- |

|  |  |  | **Depression** | **(95% CI)** | **(95% CI)** |
| --- | --- | --- | --- | --- | --- |
| Gestational week | <12 weeks | 30 | 26 | 1 | 1 |
|  | 12-28 weeks | 63 | 124 | 0.4 (0.2, 0.8) | 0.4 (0.2, 0.8) |
|  | 29-40 weeks | 43 | 64 | 0.6 (0.3, 1.1) | 0.6 (0.3, 1.2) |
| Para | Nulliparous | 40 | 95 | 0.5 (0.3, 0.8) | 0.9 (0.5, 1.5) |
|  | Multiparous | 96 | 119 | 1 | 1 |
| Type of pregnancy | Planned | 88 | 171 | 1 | 1 |
|  | Unplanned | 48 | 43 | 2.2 (1.4, 3.6) | 1.8 (1, 3.1) |
| Relationship with husband | Good | 55 | 102 | 1 | 1 |
|  | Moderate | 69 | 107 | 1.2 (0.8, 1.9) | 1.2 (0.7, 1.9) |
|  | Poor | 12 | 5 | 4.5 (1.3, 13.3) | 1.6 (0.3, 8.9) |
| Relationship with in-laws | Good | 91 | 178 | 0.3 (0.06, 2.08) | 2.6 (0.2, 38.7) |
|  | Moderate | 27 | 28 | 0.6 (0.1, 4) | 4.7 (0.3, 71.7) |
|  | Poor | 15 | 6 | 1.7 (0.2, 12.6) | 11.2 (0.6, 194) |
|  | Dead/ | 3 | 2 | 1 | 1 |
|  | Separated |  |  |  |  |
| Domestic violence | Yes | 11 | 7 | 2.6 (1, 6.9) | 1.2 (0.3, 5.7) |
|  | No | 125 | 207 | 1 | 1 |
| Sexual violence | Yes | 13 | 6 | 3.7 (1.4, 9.9) | 3.3 (1.1, 9.6) |
|  | No | 123 | 208 | 1 | 1 |
| Polygamous husband | Yes | 8 | 1 | 13.3 (1.6, 107.7) | 13.6 (1.1, 163) |
|  | No | 128 | 213 | 1 | 1 |
| Previous disease | Yes | 22 | 15 | 2.6 (1.3, 5.1) | 2.4 (1.1, 5.3) |
|  | No | 114 | 199 | 1 | 1 |

Gestational weeks, intimate partner violence, unwanted pregnancy, a history of previous disease, and multiple married husbands were found to be significantly associated with the development of depression in pregnancy. Those who were in their 2nd trimester of pregnancy had 60% less chance of developing antenatal depression than those who were in the first trimester. (AOR: 0.4, 95% CI: 0.2, 0.8) Table 5.

**Table 5.** Multivariate logistic regression of possible factors associated with antenatal depression in women visiting for antenatal check-ups in UHC, Lohagara, and Khan General Hospital, Lahuria from July to August 2023.

**General Hospital, Lahuria from July to August 2023.**
| Variables | Category | Reference | Odds | P>\|z\| |
| --- | --- | --- | --- | --- |

|  |  | Category | ratio |  |
| --- | --- | --- | --- | --- |
| Gestational week | 13-28 | <13 | 0.40967 | **0.008** |
|  | >28 |  | 0.59227 | 0.15 |
| Para | Nulliparous | Multiparous | 0.88095 | 0.633 |
| Type of pregnancy | Unplanned | Planned | 1.81135 | **0.027** |
| Relationship with husband | Moderate | Good | 1.15832 | 0.562 |
|  | Poor |  | 1.59752 | 0.593 |
| Relationship with in-laws | Good | Dead/ Separated | 1.85478 | 0.653 |
|  | Moderate |  | 3.57372 | 0.363 |
|  | Poor |  | 8.56358 | 0.098 |
| Domestic violence | Yes | No | 1.2175 | 0.803 |
| Sexual violence | Yes | No | 3.09672 | **0.03** |
| Polygamous husband | Yes | No | 13.6921 | **0.04** |
| History of previous diseases | Yes | No | 2.56009 | **0.025** |

Among the pregnant women suffering from depression, 53.6% (30) were in the first trimester. 33.7% (60) of prenatally depressed women were between 13 to 28 weeks of gestation. 40% (43) were passing their 3^rd^ trimester of pregnancies.

The odds ratio revealed three times increase in expectant women who had a history of experiencing intimate partner violence than those who did not have (AOR: 3.3, 95% CI: 1.1, 9.7).

Women facing intimate partner violence showed elevated levels of various forms of antenatal depression. Roughly 21% (4) of pregnant women with a background of intimate partner violence did not experience depression or had minimal symptoms, 47.4% (9) had mild depression, 15.8% (3) had moderate depression, and another 15.8% (3) dealt with severe depression.

Type of pregnancy was also an important contributing factor as the odds ratio raised about two times in cases of unplanned pregnancy (AOR: 1.8, 95% CI: 1, 3).

women had a history of unplanned pregnancy. Among them, 52.8% (48) were experiencing prenatal depression. 34% (98) of the total 259 expectant mothers having a history of intended pregnancies, were suffering from antenatal depression.

The incidence of moderate depression was nearly identical in both intended and unintended pregnancies, ranging from 13.2% to 13.5%. However, in the case of unwanted pregnancies, the rates of mild and severe depression were higher (mild depression: 48.4%, severe depression: 8.8%). Out of the 91 women with a history of unintended pregnancies, 48.4% (44) reported mild depression, 13.2% (12) experienced moderate depression, and 8.8% (8) had severe depression. Additionally, 29.7% (27) showed no or minimal signs of depression.

In contrast, among the total of 259 women with planned pregnancies, 37.5% (97) were dealing with mild depression, 13.5% (35) reported moderate depression, and 1.5% (4) experienced severe depression. Moreover, 47.5% (123) exhibited no or minimal signs of depression.

Women suffering from a disease from the past showed a 2 times escalated odds ratio than those who were free from any medical condition (AOR: 2.4, 95% CI: 1.1, 5.2).

Out of the pregnant women undergoing prenatal depression, 59.5% (22) had a history of prior diseases or illnesses. Within this group, 48.7% (18) were dealing with mild depression, 19% (7) with moderate depression, and 8.1% (3) with severe depression. Additionally, 24% (9) showed no or minimal signs of depression.

On the other hand, pregnant women married to husbands who have multiple wives unveiled a 13 times higher odds ratio than those having single-married husbands (AOR: 13.6, 95% CI: 1.1, 164).

Approximately 89% (8) of women with a history of multiple-married husbands were grappling with prenatal depression. Within this group, approximately 55.6% (5) were undergoing mild to moderate depression, while the remaining 33.3% (3) were dealing with severe depression during pregnancy. The rest 11.1% did not experience depression or had minimal symptoms.

## Discussion

This study aimed to assess the prevalence and evaluate the sociodemographic, obstetric, psychosocial, psychological, and disease and treatment-related determinants associated with the development of antenatal depression in a rural subdistrict in Narail. The point prevalence of antenatal depression accounted for 39% (38.86%, 95% confidence interval: 33.9% to 44%) in this study. The elevated odds ratio of Antenatal Depression Syndrome (ADS) is attributed to several significant social determinants, including sexual violence, and unplanned pregnancy. Additionally, having a polygamous husband is identified as a crucial psychological factor, while a history of previous disease serves as a notable disease and treatment-related factor contributing to the increased odds of ADS. Gestational week is an important obstetric factor associated with depression in pregnancy.

The obtained prevalence of depression in pregnancy in this study is in agreement with the prevalence in lower (34.0%, 95% Confidence Interval: 33.1%-34.9%) and middle-income countries ( 22.7%, 95% Confidence Interval: 20.1%-25.2%) (20). The result of this study also aligns with the study conducted by Gausia et al. (33%, 95% CI, 27.6–37.5) (13), and Tasnim et al. (36.2% in patients with GDM) (21), and that is perhaps because of similar locations. However, this is higher than the study of Nasreen et al. (18.3%, 95% CI:15.9%-20.7%) (22). That is probably because of the differences in the research methodology like sample size and study area. This study was conducted in Narail, which is a district of Khulna division, while the study conducted by Nasreen et al. was in the Mymensingh division, and there was a significant gap between the period of conduction of both studies. The sample sizes are also different from each other. Another study was conducted in a rural district in Sylhet which showed a higher prevalence (56.6%, 95.5% CI 50.0–63.0%) of ADS than the prevalence obtained from this study. The reason behind this distinction can be the difference in the location and number of participants involved. A majority of women in the Khulna division, specifically 88.6%, have no more than a primary education. 61% of mothers are unaware of the presence of Maternal Health Clinics (MHC) in this area, and 36% of them are receivers of any form of antenatal care (ANC) only. Additionally, ANC services from government healthcare facilities are sought by 47% of expectant mothers. Regardless of potential complications and warning signs, 95% of births usually take place at home that are assisted by untrained birth attendants. Merely, postnatal care (PNC) is provided among 19.75% of mothers and 12.3% of infants. Multiple factors contribute to the limited utilization of Maternal Health Clinic (MHC) services, including poor communication, insufficient awareness of MHC services, limited financial resources, decision-making processes, and the absence of a companion for accessing healthcare services (23). On the other hand, variations in the sample sizes are also an influencing factor for the differences.

Violence is a common problem in Bangladesh and it is one of the countries having the highest records of violence (24). Intimate partner violence was revealed as a significant contributing factor in many studies which include domestic and sexual violence. It encompasses various forms of abusive behavior, including physical violence that occurs at any point in the relationship, instances of forced sexual activity, and physical violence specifically during pregnancy. (25). 37% of women living in cities and half of the village women are sufferers of lifetime sexual violence. Causes include the history of physical abuse of mothers-in-law by fathers-in-law, The degree of husband’s controlling conduct, and the occurrence of forced or coerced initial sexual activity. In rural areas, the likelihood of this violence increased when women were between the ages of 20-24 as opposed to 15-19 and when there was a dowry demand during marriage (26). In Bangladesh, a percentage of women believe, their husbands have the right to raise their hands on them. Some women, who have been seeing their mothers become dominated and being hit by their fathers for a long time, take it easy to believe that their husbands can not only dominate them but also bear the right to beat them. These two groups of women are more likely to be the victims of intimate partner violence (27). This study found intimate partner violence as a significant contributing factor to antenatal depression which is similar to the study conducted by Peltzer et al. in Thailand and Insan et al. in Bangladesh (12, 28).

This study obtained unplanned pregnancy as a responsible factor for antenatal depression. Brazilian women who have experienced unplanned pregnancies face a 2.5 times higher risk of experiencing depression during both pregnancy and the postpartum period compared to their counterparts who have had planned pregnancies (29). Postnatal depressive symptoms among socio-economically disadvantaged rural Bangladeshi women are notably linked to the perception of paternal pregnancy unwantedness and couple pregnancy discordance. Additionally, maternal intentions and pregnancy discordance are associated with prenatal depressive symptoms in this population (30). A study conducted by Surkan et al. in northwestern Bangladesh and another study conducted by Gausia et. al in eastern Bangladesh showed that unwanted pregnancy is a significantly associated factor of antenatal depression which is similar to this study (13, 30).

Within the realm of obstetric factors, the gestational week emerges as a notable element influencing the onset of prenatal depression. The study reveals a substantial decrease in the likelihood of developing depression, amounting to a 60% reduction during the second trimester compared to the first trimester. This aligns with findings from a systematic review and meta-analysis conducted by Okagbue et al., encompassing 26 articles, which underscores that the prevalence of antenatal depression tends to be lower between the 13th and 28th weeks of gestation (31).

While some studies have identified family support and a preference for the male gender within families as significant contributing factors to prenatal depression (12, 13), this particular study did not observe such associations. It suggests that with the progression of time, contemporary family members may be more attentive and supportive of pregnant mothers than in the past. Additionally, the inclination toward preferring male children within families appears to be less prevalent today compared to historical trends. This study specifically noted that 18.5% (65) of women reported a history of male baby preference from their husbands, and 18% (63) from their families. However, a substantial majority, around 82%, did not report any history of a preference for male children from either their husbands or family members.

However, unlike other studies, this study identified noteworthy connections between a previous history of any disease and having a polygamous husband with depression during pregnancy. In a study carried out by Nasreen et al. in rural sub-districts of Mymensingh, a significant correlation was identified between a previous history of depression and prenatal depression (22). This research revealed that women with a previous mental health condition, as well as those with a history of other ailments such as Diabetes mellitus, Hypertension, and Thyroid disorders, are prone to experiencing depression during pregnancy.

## Limitations

In summary, this study faced limitations primarily stemming from a small and constrained sample size due to time and resource limitations. The study was carried out with women attending antenatal check-ups at a particular government and private hospital in Lohagara. Nevertheless, there could be a subset of women who do not seek medical attention throughout their entire pregnancy unless they experience extreme physical challenges. In many instances, these women opt for home deliveries assisted by unskilled birth attendants, bypassing hospitals during childbirth. For this reason, the findings may not fully capture the diversity of the entire community population, limiting the generalizability of obtained results.

## Conclusion

In conclusion, the prevalence and associated factors of antenatal depression in rural Bangladesh highlight a critical public health concern with far-reaching consequences for both mothers and newborns. Antenatal depression is frequently observed in the rural areas of Bangladesh, emphasizing the importance of increasing awareness among healthcare professionals and family members. This awareness is crucial to offer additional mental support to pregnant women, especially in their first and third trimesters of gestation. Furthermore, formulating strategic plans and policies is essential to reduce intimate partner violence and discourage polygamy. The situation also underscores the necessity of providing extra care to mothers with a history of health-related issues and offering additional counseling to those who find themselves unexpectedly pregnant.

To effectively tackle this issue, it is imperative for the government, stakeholders, and policymakers to collaborate on comprehensive national programs and health education campaigns. Resource allocation and the formulation of definitive policies are crucial steps toward raising awareness and destigmatizing mental health concerns in rural communities. By addressing the root causes and promoting a proactive approach to mental health, we can work towards reducing the prevalence of antenatal depression and ultimately safeguarding the well-being of both mothers and newborns in rural Bangladesh.

## Data Availability

All raw data files are available in the figshare database (DOI Number: 10.6084/m9.figshare.24994110).

https://figshare.com/articles/dataset/_b_Prevalence_and_associated_factors_of_antenatal_depression_in_rural_Bangladesh_b_/24994110

## Supporting Information

S1 File. Dataset. https://doi.org/10.6084/m9.figshare.24994110 (xlsx)

## Acknowledgement

I am profoundly thankful to the Almighty for granting me the opportunity to pursue MPH (Epidemiology) at North South University. I am also grateful to North South University for allowing me to do the research for the partial fulfillment of my MPH degree. My gratitude extends to my supervisor, Dr. Dipak Kumar Mitra, Ph.D., MPH, MBBS, the current chairman of the Department of Public Health at North South University, for his kind advice and guidance. I am greatly thankful to Dr.S M Mashud, UH&FPO, Upazila Health Complex, Lohagara, for his wonderful guidance and unwavering support to carry out the study.

## Notes

### Competing Interest Statement

The authors have declared no competing interest.

### Funding Statement

This study did not receive any funding

### Author Declarations

Approved by-Institutional Review Board/Ethics Review Committee (IRB/ERC), North South University Approval Number: 2023/OR-NSU/IRB/1224

